# Lung remodeling regions in long-term Covid-19 feature basal epithelial cell reprogramming

**DOI:** 10.1101/2022.09.17.22280043

**Authors:** Kangyun Wu, Yong Zhang, Stephen R. Austin, Huqing Yin Declue, Derek E. Byers, Erika C. Crouch, Michael J. Holtzman

**Affiliations:** Pulmonary and Critical Care Medicine, Department of Medicine, Washington University School of Medicine, Saint Louis, MO 63110; Department of Pathology and Immunology, Washington University School of Medicine, Saint Louis, MO 63110; Department of Cell Biology and Physiology, Washington University School of Medicine, Saint Louis, MO 63110

## Abstract

Respiratory viruses, including SARS-CoV-2, can trigger chronic lung disease that persists and even progresses after expected clearance of infectious virus. To gain an understanding of this process, we examined a series of consecutive fatal cases of Covid-19 that came to autopsy at 27-51 d after hospital admission. In each patient, we identify a stereotyped bronchiolar-alveolar pattern of lung remodeling with basal epithelial cell hyperplasia and mucinous differentiation. Remodeling regions also feature macrophage infiltration and apoptosis and a marked depletion of alveolar type 1 and 2 epithelial cells. This entire pattern closely resembles findings from an experimental model of post-viral lung disease that requires basal-epithelial stem cell growth, immune activation, and differentiation. The present results thereby provide evidence of possible basal epithelial cell reprogramming in long-term Covid-19 as well and thereby a pathway for explaining and correcting lung dysfunction in this type of disease.

---

The Covid-19 pandemic spotlights the need to better understand both acute and chronic disease triggered by severe respiratory viral infection. The acute phase of disease with severe pneumonia and lung injury dominated the early management and study of the crisis (1). However, even then, it appeared likely that progressive and often long-term disease was also a significant cause of morbidity and mortality. Indeed, a high percentage of Covid-19 patients survived the acute infectious illness only to experience the major degree of organ dysfunction over a more prolonged time course during and after the initial hospitalization (2, 3). This outcome was consistent with previous observations that other types of respiratory viruses can also trigger a pathway to long-term immune-mediated disease. Thus, viral initiation, exacerbation, and progression of chronic lung disease can be found in clinical observations and corresponding experimental models of asthma, COPD, and related inflammatory disease phenotypes (4). Together, these observations raise the likelihood that the host response to the virus can be reprogrammed from protection to an abnormal remodeling response in susceptible individuals. Whether and how this alternative pathway might also be linked to progressive and/or long-term Covid-19 still needs to be defined.

To address this issue, we studied lung tissue samples from autopsies of Covid-19 patients to obtain cell and molecular insights in the context of previous observations. Our analysis focused on a consecutive series of Covid-19 patients that died long after onset of illness (27-51 d after hospital admission) to provide a snapshot of long-term post-viral lung disease. In fact, this opportunity for analysis of lung tissue was largely unprecedented in human studies of this disease process. In that context, we found consistent and striking basal-epithelial cell hyperplasia that extended beyond the usual airway location and instead moved into the distal airspaces of the lung. As introduced above, this remodeling process was captured long after the start of the initial illness but in close association with morbidity and mortality from the disease. Moreover, this same pattern of progression was predominant in experimental models of viral infection using natural pathogens for mice such as Sendai virus (SeV) (5-7) and for humans such as influenza A virus (IAV) and respiratory enterovirus-D68 (8, 9). Those studies ultimately identified a subset of basal-epithelial stem cells (basal-ESCs) that are critical for homeostasis but also reprogrammable to become an essential driver of lung remodeling signatures (7). Here we identify similar circuitry for lung remodeling in post-viral Covid-19 patients with the possibility that the same components can be targeted to address the problem of long-term Covid-19 in the lung and perhaps other tissue sites.

## Materials and Methods

### Human sample procurement and processing

Human lung tissue was obtained from a series of consecutive autopsies performed from April-August 2020 at Barnes-Jewish Hospital. Case histories are presented in Supplemental Text with a summary of clinical characteristics in Supplemental Table 1. As controls, human lung samples were obtained from an Advanced Lung Disease Tissue Registry that contains whole lung explants harvested but not used for lung transplantation as described previously (10, 11) and from a tissue procurement service (IIAM, Edison, NJ). All human studies were conducted with protocols approved by the Washington University Institutional Review Board.

### Tissue staining, immunohistochemistry, and analysis

Tissues were fixed with 10% formalin, embedded in paraffin, cut into 5-µm sections, and adhered to charged slides. Sections were deparaffinized in Fisherbrand® CitroSolv® (Fisher), hydrated, and treated with heat-activated antigen unmasking solution (Vector Laboratories, Inc). Tissue processing and then hematoxylin-eosin, PAS-hematoxylin, and Gomori trichrome staining was performed as described previously (6, 8). Immunostaining was performed using the following primary antibodies: rabbit anti-ACE-2 pAb (ab65863, Abcam), rabbit anti-KRT5 pAb (ab53121, Abcam) and mAb (clone EP1601Y, ab52635, Abcam), rabbit anti-AQP3 pAb (ab125219, Abcam), mouse anti-CD68 mAb (clone Kp-1, Sigma-Aldrich), mouse anti-MUC5AC mAb (clone 45M1, Thermo Scientific and Santa Cruz Biotechnology), mouse anti-HT2-280 pAb (TB-27AHT2-280, Terrace Biotech), rabbit anti-surfactant protein C (SFTPC) pAb (ab90716, Abcam), rabbit anti-podoplanin (PDPN) mAb (clone EPR7072, Abcam), rabbit anti-MUC5B pAb (ab87276, Abcam), rabbit anti-Ki-67 mAb (clone D2H10, Cell Signaling), and rabbit active (cleaved) caspase-3 mAb (clone 5A1E, Cell Signaling). Antibody binding was detected with Alexa Fluor 488 or 594 conjugated secondary antibodies (Invitrogen). All sections were counterstained with DAPI and were imaged using a Leica DM5000 B microscope for conventional imaging. Staining was quantified in whole lung sections using a NanoZoomer S60 slide scanner (Hamamatsu) and ImageJ software as described previously (7). For the present experiments, an entire lung section was scanned and analyzed for each patient or subject (n=5 per group) to assess lung remodeling region phenotype.

### Statistical analysis

All data are presented as mean and s.e.m and are representative of five Covid-19 patients or five non-disease subjects. Unpaired student’s t-test as well as mixed-model repeated measures analysis of variance with Tukey correction for multiple comparisons were used to assess statistical significance between means. In all cases, significance threshold was set at *P*<0.05. The number of patients and subjects for each experimental condition is defined in the figure legends.

## Results

To understand clinical observations of long-term disease after SARS-CoV-2 infection, we developed a system to obtain and analyze the lung tissue samples from a consecutive set of hospitalized patients that underwent autopsy for Covid-19 as a primary cause of death. This group of five patients was highly relevant to long-term post-viral disease since they died at 27-51 d after initial presentation for care and positive testing for SARS-CoV-2 infection (Supplemental Table 1). Examination of lung sections showed similar expression of the SARS-CoV-2 receptor ACE-2 on airway and alveolar epithelial cells in patients and non-disease control subjects (Fig. 1a), suggesting viral capacity for primary infection of host epithelial cells in proximal and distal sites. In concert with this possibility, hematoxylin-eosin staining showed alveolar epithelial disruption in all patients consistent with diffuse organizing alveolar damage (Fig. 1b). In addition, this staining revealed a stereotyped pattern of airway epithelial cell hyperplasia that extended into distal airspaces that were filled with cellular and mucinous material and were adjacent to immune cell accumulation in blood vessels and tissue (Fig. 1b). PAS-hematoxylin staining confirmed these findings including accumulation of apparent mucus in these same locations (Fig. 1b). Gomori trichrome staining indicated that these same bronchiolar-alveolar lung remodeling regions were also sites for collagen production consistent with a fibrotic reaction (Fig. 1b).

**Figure 1.**
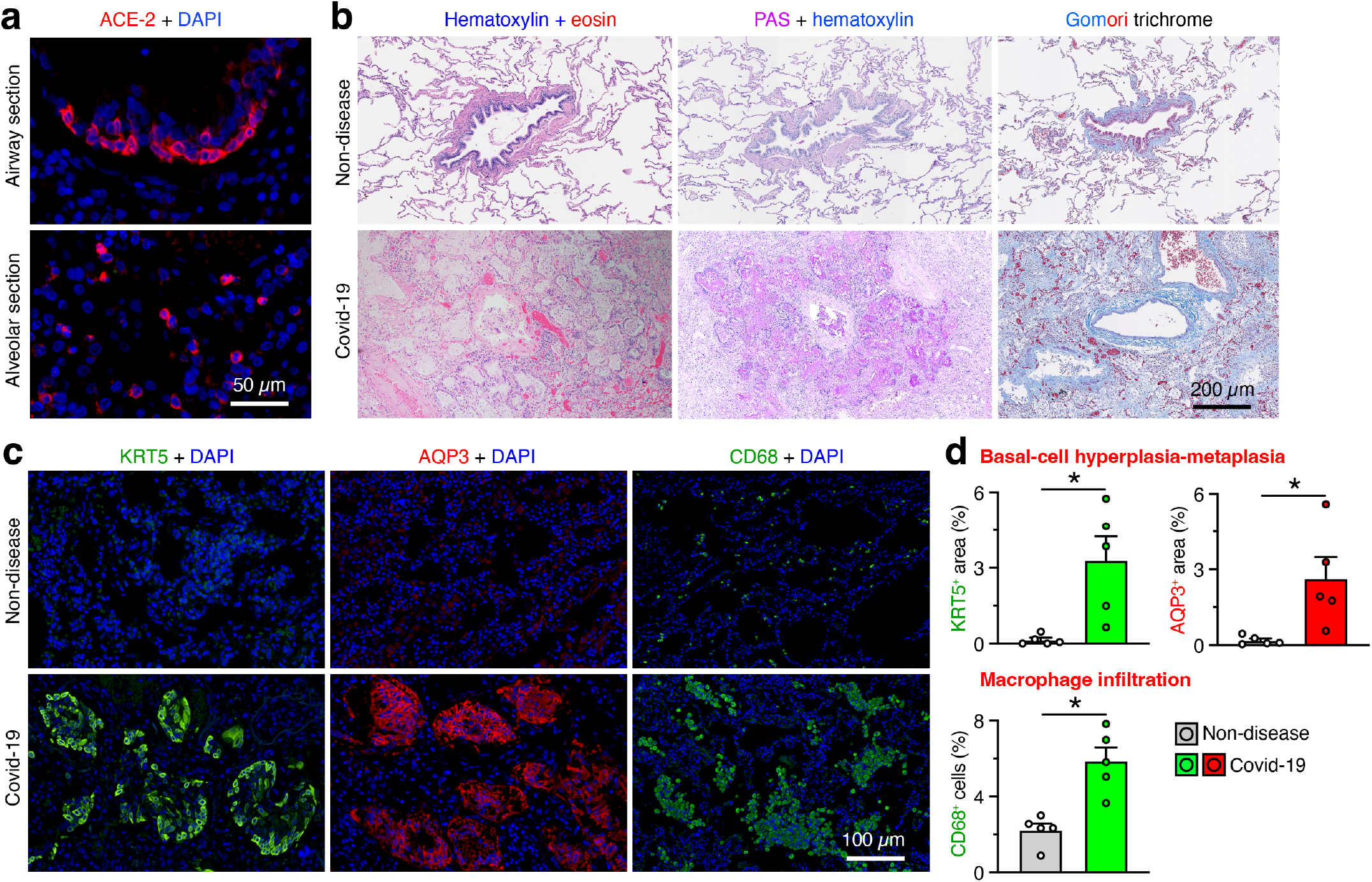
Basal epithelial cell hyperplasia-metaplasia and mucinous differentiation along with fibrosis and macrophage infiltration in lung remodeling regions in Covid-19 patients. **a**, Immunostaining for ACE2 with counterstaining for DAPI in airway and alveolar sections of representative lung tissue from Covid-19 patients. Scale bar=50 µm. **b**, Hematoxylin-eosin, PAS-hematoxylin, and Gomori trichrome staining of lung sections from Covid-19 patients and non-disease controls. For trichrome staining: collagen/mucus blue, cytoplasm/keratin/muscle red, nuclei black. Scale bar=200 µm. **c**, Immunostaining for KRT5, AQP3, and CD68 with DAPI counterstaining in lung sections from Covid-19 patients and non-disease control subjects. Scale bar=100 µm. **d**, Quantitation of staining for conditions in (c). Data are representative of 5 patients (at 27-51 d after hospital admission for Covid-19) and 5 control subjects per staining condition. Values represent mean and s.e.m; \**P*<0.05 (n=5 patients or subjects per group).

Immunostaining with the basal epithelial cell markers KRT5 and AQP3 (7) more specifically identified the presence of basal-epithelial cell clusters extending into former alveolar spaces (Fig. 1c). These remodeling sites also showed increased staining for macrophage surface receptor CD68, compared to non-disease controls (Fig. 1c). Quantitative levels of KRT5^+^, AQP3^+^, and CD68^+^ staining were each significantly increased in Covid-19 patients compared to non-disease subjects (Fig. 1d). This result served as an index of basal-epithelial cell hyperplasia-metaplasia and macrophage infiltration to mark the extent of lung remodeling regions in Covid-19 patients.

To better define the observed damage to the alveolar epithelium, we also checked the spatial distribution and morphology of alveolar type 2 (AT2) and type 1 (AT1) cells during late-stage disease. As expected, immunostaining showed that KRT5^+^ basal cells were restricted to airway mucosal sites and HT2-280^+^SFTPC^+^ AT2 cells and PDPN^+^ AT1 cells were confined to alveolar sites in non-disease control subjects (Fig. 2a,b). In contrast, KRT5^+^ cells were abundant while HT2-280^+^, SFTPC^+^ and PDPN^+^ cells were nearly absent in the lung remodeling regions of Covid-19 patients (Fig. 2a,b).

**Figure 2.**
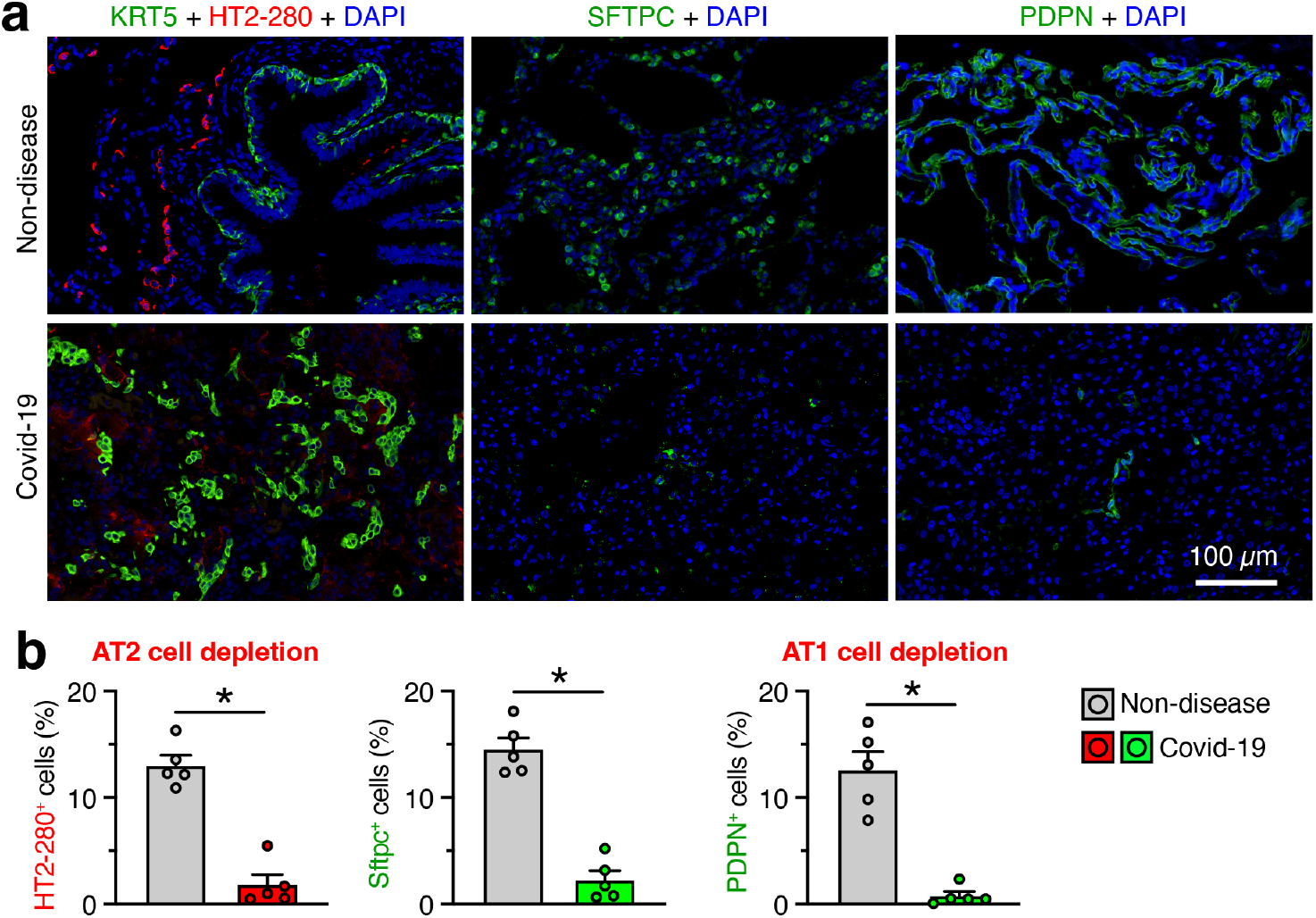
Alveolar epithelial cell loss in lung remodeling regions in Covid-19. **a**, Immunostaining for KRT5 plus HT2-280, SFTPC, and PDPN with DAPI counterstaining in lung sections from Covid-19 patients and non-disease controls. Scale bar=100 µm. **b**, Quantitation of staining for conditions in (b). Data are representative of 5 patients and 5 control subjects per staining condition. Values represent mean and s.e.m; \**P*<0.05 (n=5 patients or subjects per group).

As a further readout of post-viral lung remodeling, we also characterized mucinous differentiation as defined by expression of the predominant lung gel-forming mucins MUC5AC and MUC5B. In non-disease control subjects, we found the usual pattern of mixed MUC5AC^+^ and MUC5B^+^ staining in airway mucosal epithelium, predominant MUC5B^+^ staining in submucosal glands, and no detectable MUC5AC^+^ or MUC5B^+^ staining in alveolar epithelium (Fig. 3a). As noted previously (6), non-disease control subjects (that were on mechanical ventilation similarly to patients) exhibited variable amounts of airway mucus staining, but none of the controls showed lung remodeling regions as found in Covid-19 patients. In contrast, we detected mixed MUC5AC^+^ and MUC5B^+^ staining that was generally co-localized to mucous cells in lung remodeling regions for each of the Covid-19 patients (Fig. 3b). Quantitation of mucin staining showed a significant increase in MUC5AC^+^ and MUC5B^+^ staining at these sites compared to comparable bronchiolar-alveolar sites in non-disease controls (Fig. 3c).

**Figure 3.**
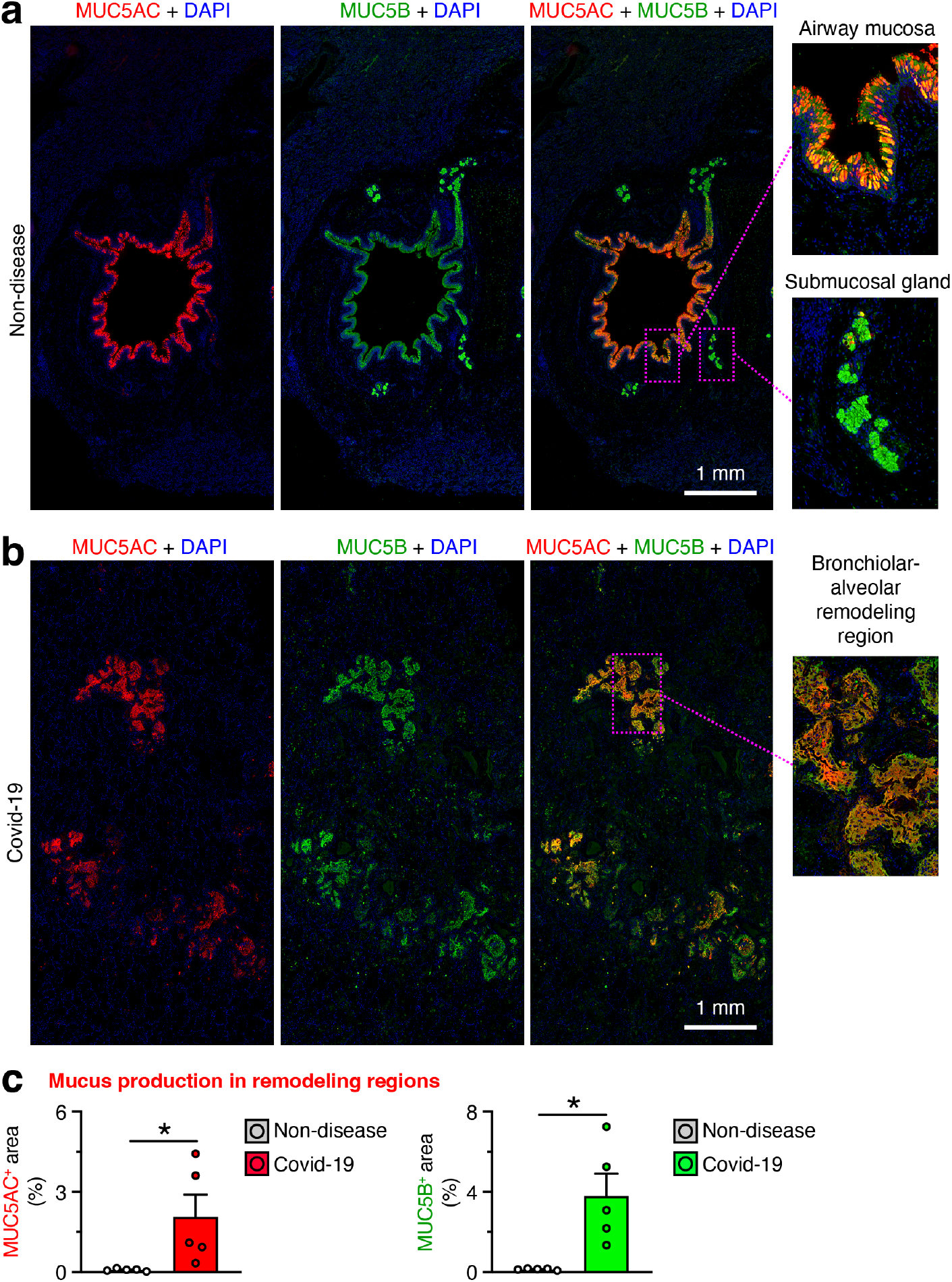
MUC5AC with MUC5B expression in lung remodeling regions in Covid-19. **a**, Immunostaining for MUC5AC and MUC5B with DAPI counterstaining in lung sections from non-disease control subjects. Scale bar=1 mm; Original magnification, x5 (insets). **b**, Immunostaining for MUC5AC and MUC5B with DAPI counterstaining in lung sections from Covid-19 patients. Scale bar=1 mm; Original magnification, x3 (insets). **c**, Quantitation of staining for conditions in (b). Data are representative of 5 patients per staining condition. Values represent mean and s.e.m; \**P*<0.05 (n=5 patients or subjects per group).

We also aimed to better define the net increases in basal epithelial cells and macrophages as an index of epithelial and immune cell activation. We found a modest but significant increase in Ki-67^+^ cells as a sign of cell proliferation along with a more marked increase in active caspase-3^+^ cells as a marker of programmed cell death (apoptosis) (Fig. 4a,b). Moreover, the active caspase-3 signal was not localized to the decreased population of HT2-280^+^ AT2 cells but instead was co-localized primarily to the increased population of CD68^+^ lung macrophages (Fig. 4c). Together, these findings were consistent with macrophage infiltration and accumulation despite an increased level of programmed cell death even at this late stage of post-viral lung disease.

**Figure 4.**
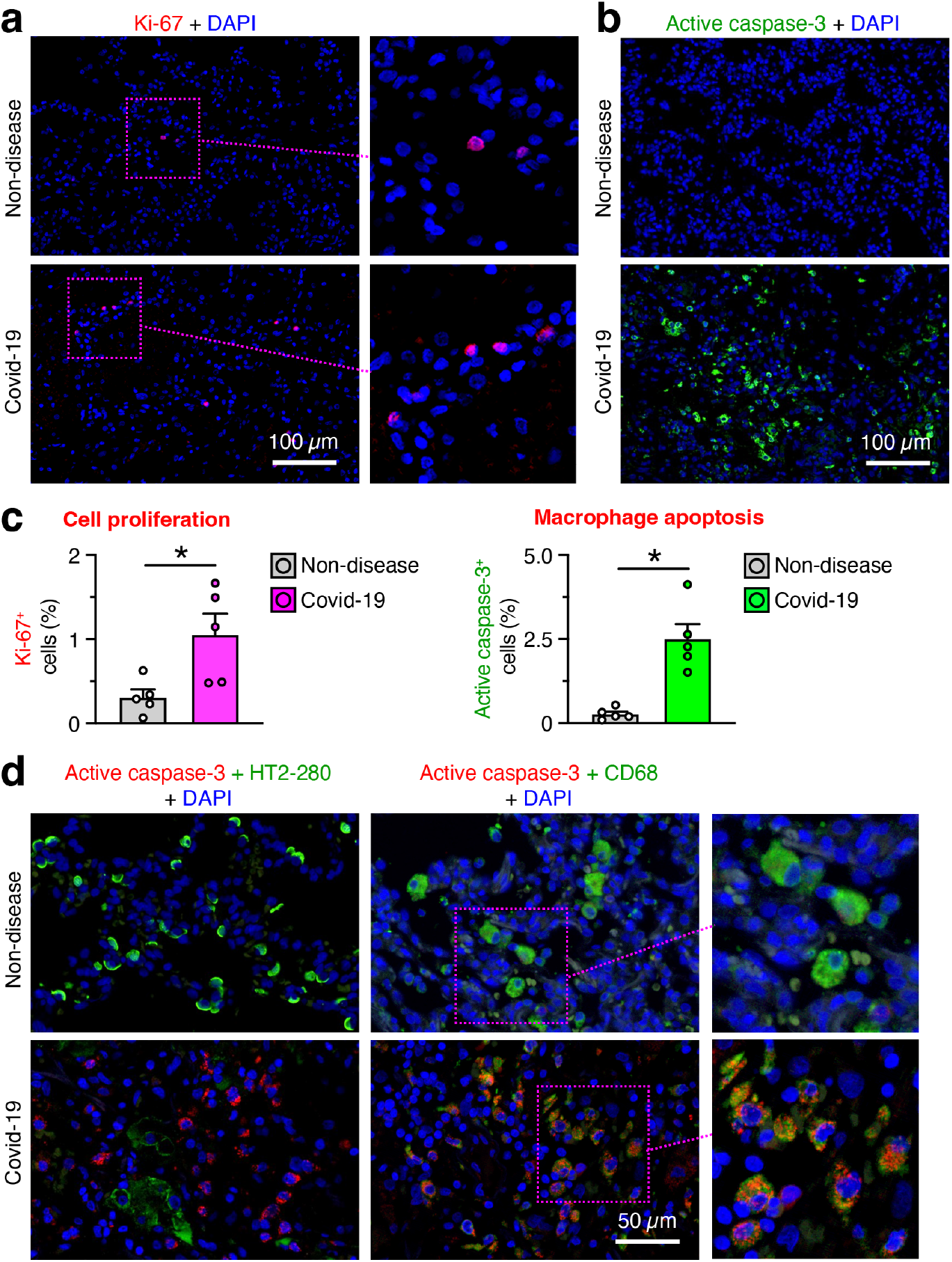
Cell proliferation and apoptosis in lung remodeling regions in Covid-19. **a**, Immunostaining for Ki-67 with DAPI counterstaining in lung sections from Covid-19 patients and non-disease controls. Scale bars=400 µm; Original magnification, x5 (inset). **b**, Immunostaining for active caspase-3 with DAPI counterstaining in lung sections for groups in (a). Scale bars=100 µm. **c**, Quantitation of staining for conditions in (a,b). **d**, Immunostaining for active-caspase-3 plus HT2-280 or CD68 with DAPI counterstaining for groups in (a). Scale bar=50 µm; Original magnification, x3 (inset). Data are representative of 5 patients and 5 control subjects per staining condition. Values represent mean and s.e.m; \**P*<0.05 (n=5 patients or subjects per group).

## Discussion

This study analyzed a series of autopsies from Covid-19 patients with late-stage lung disease at least 27-51 d after initial viral infection. The results reveal a bronchiolar-alveolar lung remodeling process that is characterized by: (1) basal epithelial cell hyperplasia and metaplasia with extension into former alveolar spaces; (2) co-localized depletion of alveolar types of epithelial cells normally found in these spaces; (3) epithelial stem-progenitor cell differentiation to mucous cells with mucins characteristic of mucosal and submucosal locations; and (4) closely associated lung macrophage infiltration. Here we place our findings in the context of human clinical studies and comparable animal models.

Relative to human studies, there is only limited previous data on post-viral lung remodeling. The Covid-19 pandemic created an unprecedented opportunity to define this process in vivo. Initial data from Covid-19 patients documented the early phase of diffuse alveolar damage typical of lung injury and acute respiratory distress syndrome (ARDS) (12-14). Thereafter came reports of basal-epithelial cell repair (15, 16), AT2 cell loss (17), macrophage accumulation (18, 19), predominant MUC5AC^+^ mucus production (20), and selectively MUC5B^+^ microcysts (21) at subsequent stages of disease in lungs of Covid-19 patients. The present series of entirely long-term cases was further characterized by the consistent development of lung remodeling regions with KRT5^+^-AQP3^+^ basal-epithelial cell hyperplasia-metaplasia, combined AT1-AT2 cell dropout, mixed MUC5AC^+^-MUC5B^+^ mucus production, and CD68^+^ macrophage accumulation despite accelerated caspase-3^+^ apoptotic turnover. Together, the data provide histological, cellular, and molecular definition of the switch from acute lung injury to chronic bronchiolization that was suggested in descriptive reports of autopsies after presumed viral infection (22).

These new findings also enable comparison of the present data to animal models to better define the pathogenesis of post-viral lung disease. At present, experimental models of SARS-CoV-2 infection do not yet provide a model of long-term lung disease with high fidelity to the human phenotype noted above. This discrepancy is likely a reflection of the inability to duplicate the usual pattern of severe respiratory infection. In that regard, the top-down spread and consequent severity of infection can be achieved with the natural Sendai virus (SeV) pathogen in mice. This type of infection thereby yields a remarkably similar pattern of acute infectious illness and in turn manifests the key signatures of post-viral lung remodeling disease found in Covid-19 (7, 9, 23). This experimental model also reveals a basal-ESC subset that jumps the usual bronchiolar-alveolar boundary and grows into distal airspaces as a new site for differentiation and immune activation that is required for long-term dysfunction (7). Single-cell analysis marks this cell subset and its descendent lineage with macrophage chemokine *Cxcl17* expression in mice (7) and humans (https://www.proteinatlas.org). Further, *CXCL17* mRNA is increased markedly in bronchoalveolar lavage (BAL) samples from Covid-19 patients (24). Whether CXCL17 is detectable in long-term disease or functions as a downstream effector signal for macrophage participation still needs to be defined. Nonetheless, macrophage participation is essential for long-term disease in the SeV mouse model (5, 23, 25) consistent with macrophage infiltration found in remodeling regions of long-term Covid-19 patients. A similar post-viral process can be detected after severe infection with influenza A virus in mice (8) and even in milder infection with enterovirus (EV-D68) in mice engineered for deficiency in airway epithelial interferon-signaling (9). A similar type 1 interferon deficiency is associated with increased severity in Covid-19 (26), consistent with observations that severity of illness is linked to long-term disease in mouse models and humans (3, 8, 9).

Together, the present data for Covid-19 patients fit a consistent paradigm for long-term post-viral lung disease found in natural and experimental settings. Our perspective is that epithelial barriers maintain a basal-ESC subset that is tasked for repair and regeneration after injury, including the damage from severe viral infection. However, this same system can orchestrate a tissue remodeling process that is pathogenic since basal epithelial cell depletion can attenuate post-viral disease in the lung and disrupt homeostasis at other barrier sites (7). The presently identified markers for the relevant basal-ESC subset thereby suggest an approach to disease modification via correction of basal-ESC reprogramming. Suitable targets already exist in the form of IL-33-nucleokine and MAPK13-stress-kinase activation that allow for basal-ESC growth and mucinous differentiation in post-viral lung remodeling disease (7, 27) and perhaps other virus-triggered lung diseases (4). The present study provides a basis to define these and related strategies in lung remodeling disease due to Covid-19.

## Data Availability

All data produced in the present work are contained in the manuscript.

## Author contributions

K.W. designed and performed experiments, H.Y.D performed experiments; Y.Z. assisted with data analysis; S.R.A. and D.E.B. obtained and analyzed clinical data; E.C.C. identified autopsy cases and assisted with histopathology analysis, and M. J. H. directed the project and wrote the manuscript. All authors acquired and analyzed data.

## Acknowledgments

We thank the Pulmonary Morphology Core and the Anatomic and Molecular Pathology Core for technical support. This work was supported by grants from the National Institutes of Health (National Heart, Lung, and Blood Institute R35-HL145242, National Institute of Allergy and Infectious Diseases R01-AI130591), Department of Defense (PR190726 and PR211069), and the Harrington Discovery Institute, Cystic Fibrosis Foundation, Bebermeyer Fund, Hardy Trust, and Schaefer Fund.

