## Supplemental Text and Table 1 for "Lung remodeling regions in long-term Covid-19 feature basal epithelial cell reprogramming"

*Case histories*

### Patient 1

Patient 1 was an African American, non-Hispanic woman with a past medical history of dementia, hypertension, alcohol abuse, asthma, and chronic obstructive pulmonary disease (COPD). She presented to the Emergency Department (ED) with symptoms of atypical pneumonia and altered mental status. Patient 1 was immediately admitted to the ICU, where the patient stayed for 33 d until their death. During hospitalization, the patient was treated with glucocorticoid and hydroxychloroquine and was maintained on respiratory support that was eventually weaned to room air. However, she later became hypoxemic again requiring nasal high flow oxygen therapy Chest x-ray showed bilateral reticular opacities and patchy bibasilar, perihilar (right greater than left) opacities with interval improvement. She subsequently had further worsening of respiratory symptoms and was transitioned to comfort care before dying. An autopsy was requested by the patient's family and performed 1 d after death. The autopsy determined the cause of death to be respiratory failure secondary to severe pneumonia in a setting of SARS-CoV2 infection and underlying COPD.

### Patient 2

Patient 2 was a White, Hispanic man whose comorbidities included nonalcoholic fatty liver diseases, alcohol use, cocaine use, tobacco smoking, and obesity. He presented to the ED initially asymptomatic after a positive test for SARS-CoV-2 following exposure to a SARS-CoV-2-positive family member. The patient developed fever, malaise, anosmia, and diarrhea 3 d later, and then worsening dyspnea and cough 6 d later that required emergency medical assistance. The emergency medical service team found the patient hypoxemic ($SpO_2$ 50%). Patient 2 was admitted and intubated on the same day. The patient died 25 d later (31 d after their onset of symptoms). During his hospital stay, the patient was supported initially with Bilevel Positive Airway Pressure (BiPAP). A chest CT scan supported a diagnosis of SARS-CoV-2 pneumonia without pulmonary emboli. The patient became septic and started on empiric antibiotics, mechanical ventilation, and treatment with dexamethasone and remdesivir but did not improve over following weeks. Sedation provided for comfort after escalation of care was declined by his family. The patient developed renal failure with uremia and hyperkalemia in the context of worsening respiratory function. An autopsy was performed 3 d after death and found the primary cause of death to be acute lung injury in a fibroproliferative phase with marked alteration in lung architecture with consolidation, focal hyalin membranes, hemorrhage, bilateral pleural serosanguinous effusions with a history of acute respiratory failure secondary to SARS-CoV-2 infection. Contributory causes of death included cirrhosis with obesity and past cocaine and tobacco use.

**Patient 3**

Patient 3 was an African American, non-Hispanic man who had a past medical history of hypertension. This patient presented to the ED with a 3-d history of dry cough and malaise after Covid-19 exposure to a family member. This patient tested virus-positive and initially was sent home for monitoring. He returned to the ED 3 d later with worsening shortness of breath, thus 6 d after onset of symptoms. The patient was admitted to the hospital and intubated 4 d later. A chest CT scan showed multiple patchy areas of peripheral ground-glass consolidations throughout both lungs. He was treated with remdesivir and dexamethasone. Patient acutely decompensated and required emergent and difficult intubation with subsequent ventricular fibrillation, cardiac arrest, and resuscitation. His EEG showed profound slowing and extensive diffusion restriction on MRI. The patient also developed acute kidney injury requiring hemodialysis and *Staphylococcus aureus* and *Enterococcus faecalis* infection of his dialysis catheter requiring antibiotic treatment. The patient also experienced bradycardic arrest during dialysis and was resuscitated. The family elected to designate DNR status, and soon thereafter the patient became hypoxemic with cardiac arrest after an endotracheal tube leak An autopsy was performed 2 d after death. The primary cause of death was determined to be acute lung injury in the background of organizing phase of diffuse alveolar damage in the setting of SARS-CoV-2 infection. Contributory causes of death included obesity, hypertension, cardiomegaly, anoxic brain injury, and acute kidney injury.

**Patient 4**

Patient 4 was an African American, non-Hispanic female with pre-existing comorbidities of ischemic cardiomyopathy, coronary artery disease, COPD, obstructive sleep apnea, end-stage renal disease, and type II diabetes mellitus. Patient 4 presented to the ED with symptoms of heart failure exacerbation and tested positive for SARS-CoV-2 despite being asymptomatic. Following discharge for at-home SARS-CoV-2 monitoring, the patient developed chest pain, fatigue, and progressive shortness of breath and was re-admitted 1 d after discharge. After 13 d in the hospital, Patient 4 worsened and required intubation and ventilation and ultimately died 15 d later. Her clinical course was significant for worsening respiratory status with increasing oxygen requirements and persistent hypoxia despite maximal ventilator support, profound hypotension requiring multiple vasopressors, and worsening end organ failure. Chest imaging demonstrated worsening interstitial opacities consistent with diffuse alveolar damage. Prior to death, Patient 5 went into pulseless electrical activity arrest requiring resuscitation. Shortly thereafter, the patient went into asystolic arrest and did not recover despite resuscitation efforts. An autopsy was performed 2 d after death. The primary cause of death was determined to be diffuse alveolar damage in the setting of SARS-CoV2 infection leading to acute hypoxemic respiratory failure. Contributory causes of death include comorbidities including heart failure with ischemic cardiomyopathy, coronary artery disease, COPD, obstructive sleep apnea, end stage renal disease, and type II diabetes mellitus

**Patient 5**

Patient 5 was an African American, non-Hispanic male with a past medical history of morbid obesity, type II diabetes mellitus, hyperlipidemia, stage 3 chronic kidney disease, hypertension, and obstructive sleep apnea. The patient presented to the ED with shortness of breath, recently positive SARS-CoV-2 test, decreased $SpO_2$, and chest X-ray suggesting pneumonia. He was transferred to the ICU for BiPAP support and received treatment with tocilizumab, remdesivir, and dexamethasone along with azithromycin and rocephin. Despite treatment, his oxygen requirements increased, and he required intubation and mechanical ventilation. He also required prone position and paralysis for ventilation, inhaled nitric oxide, and transient dialysis for hyperkalemia. A week later, he also developed metabolic acidosis and hypoxemia requiring ECMO treatment. His tracheal aspirate continued to be SARS-CoV-2 positive, and blood cultures were positive for *Candida auris*. At 41 d after admission, the patient was placed on comfort care, and 10 d later, the patient died. An autopsy was performed 2 d after death. The primary cause of death was determined to be organizing alveolar damage and pneumonia consistent with Covid-19. Contributory causes of death included *Candida auris* fungemia, positive tracheal aspirate for *Klebsiella*, morbid obesity, type II diabetes mellitus, chronic kidney disease, hyperlipidemia, and hypertension.

**Supplemental Table 1. Clinical characteristics of Covid-19 patients (n=5).**

| Case number | Gender | Race | Co-morbidities | Time from diagnosis to death (d) | Cause of death |
|---|---|---|---|---|---|
| 1 | F | Black | Dementia, hypertension, alcohol abuse, asthma-COPD | 33 | Primary: respiratory failure due to Covid-19 and underlying asthma-COPD<br><br>Contributory: cardiac arrest |
| 2 | M | White | Nonalcoholic fatty liver disease, alcohol use,<br>cocaine use, tobacco smoking, obesity | 34 | Primary: Respiratory failure due to Covid-19<br><br>Contributory: cirrhosis, obesity. |
| 3 | M | Black | Hypertension | 27 | Primary: respiratory failure due to Covid-19<br><br>Contributory: obesity, hypertension, cardiomegaly, anoxic brain injury, acute kidney injury |
| 4 | F | Black | Ischemic cardiomyopathy, heart failure, COPD,<br>Obstructive sleep apnea,<br>End-stage renal disease,<br>Type II diabetes mellitus | 31 | Primary: respiratory failure due to Covid-19.<br><br>Contributory: multiple co-morbidities |
| 5 | M | Black | Obesity, diabetes, hypertension, chronic kidney disease, obstructive sleep apnea. | 51 | Primary: respiratory failure due to Covid-19.<br><br>Contributory obesity, diabetes, chronic kidney disease, hypertension, hyperlipidemia. |
